# Combining Abdominal Ultrasound and Radiography for Surgical Risk Prediction in Necrotising Enterocolitis: A Prospective Cohort Study

**DOI:** 10.1101/2025.11.03.25339365

**Authors:** Archana Priyadarshi, Rajeshwar Reddy Angiti, Shilpi Chabra, Annabel Webb, Ryan M. McAdams, Nadia Badawi, Murray Hinder, Mark Tracy

## Abstract

**Objective:** To evaluate whether combining abdominal ultrasound with radiography improves diagnostic accuracy and surgical risk prediction in neonates with suspected necrotising enterocolitis (NEC) compared with radiography alone.

**Design, setting, patients:** Prospective cohort study conducted in two tertiary neonatal intensive care units. Sixty-seven neonates with suspected NEC underwent concurrent abdominal radiography and ultrasound assessments. Imaging studies were independently reviewed by masked investigators using pre-specified criteria to classify each study as reassuring or non-reassuring.

**Main outcome measures:** The primary outcome was the need for surgical intervention. Imaging data were analysed using unsupervised k-means clustering (k = 2); logistic regression - testing associations with surgery and Principal component analysis (PCA) - to identify imaging features most contributing to group separation.

**Results:** Ultrasounds were reassuring in all cases subsequently diagnosed with non-NEC, i.e feeding intolerance, whereas most radiographs in this group were non-reassuring. Clustering based on radiographs alone did not significantly discriminate surgical risk (58.8% vs 39.4%; p = 0.11). Combined model (radiograph + ultrasound) produced two distinct clusters with significantly different surgical rates (78.3% vs 34.1%; OR 6.96, 95% CI 2.29–24.58). PCA highlighted complex ascites, absent peristalsis, and abnormal bowel perfusion as key discriminating features.

**Conclusion:** In suspected NEC, combining ultrasound with radiography significantly improves surgical risk stratification compared with radiography alone. A reassuring ultrasound reliably identified infants with feeding intolerance, suggesting potential to reduce unnecessary transfers and treatments. Larger multicentre studies are needed to validate these findings and inform development of a unified multimodal imaging score for NEC diagnosis.

**Key messages:** *What is already known on this topic:*

- NEC is an acquired intestinal disease mainly affecting preterm infants with the potential risk of serious life-long co-morbidities.
- Plain abdominal radiography remains the standard initial investigation for suspected NEC but provides limited information on bowel perfusion and viability.
- Emerging evidence supports the role of bowel ultrasound in NEC, in assessing bowel wall thickness, perfusion, peristalsis, and intra-abdominal fluid collections.

*What this study adds:*

- Using a multi-modal imaging approach (combining ultrasound and radiography) improved surgical risk stratification in neonates with suspected NEC compared with using radiography alone.
- A reassuring ultrasound may identify benign conditions initially suspected as early NEC, supporting earlier de-escalation of care.
- The study findings lay the foundation for a standardised imaging-based score combining radiograph and ultrasound findings in cases with suspected NEC, to better guide diagnosis and predict the need for surgical intervention.

## Introduction

Necrotising enterocolitis (NEC) is a severe intestinal disorder of early infancy that predominantly affects preterm infants.^1^ It involves inflammation, ischaemia, and varying degrees of bowel wall necrosis.^2^ Mortality remains high at 20-30% and up to 40% of survivors require surgical intervention, often resulting in short bowel syndrome and adverse neurodevelopmental outcomes.^3^ NEC typically affects the bowel wall. In severe disease, the bowel wall becomes ischaemic, thin, and susceptible to perforation.^4^ Milder cases may resolve completely or heal with stricture formation, whereas progressive disease can lead to peritonitis, sepsis, intra-abdominal adhesions, and haemodynamic collapse.^5^

Plain abdominal radiography remains the first-line investigation for suspected NEC.^6^ The presence of pneumatosis intestinalis or portal venous gas on the plain abdominal radiograph has traditionally defined the diagnosis of NEC, although the diagnostic exclusivity of these findings has recently been questioned.^7^ In the absence of a reliable biomarker, diagnosis often depends on clinical judgment and imaging interpretation.^8^ Because the disease can progress rapidly and the consequences of a missed diagnosis are severe, NEC is more frequently suspected than confirmed.^1^

Ultrasound (US) provides a complementary, radiation-free modality that allows bedside assessment of bowel wall thickness, vascularity, and peristalsis, which are critical features to assess for the presence of NEC disease and its progression.^4^ Radiologists have detailed the characteristic sonographic findings in NEC.^9–11^ In Australia and New Zealand neonatal clinicians routinely use point-of-care US to evaluate heart, brain and lungs.^12–14^ There is growing interest in extending this capability to abdominal ultrasound imaging, with increasing adoption of point-of-care US for early detection and monitoring of NEC.^15,16^

This study aimed to evaluate whether combining abdominal ultrasound and radiography as a multimodal imaging approach, improves diagnostic precision and surgical risk stratification in neonates with suspected NEC. We hypothesised that integrating both modalities, compared with using diagnostic radiography alone, would better identify neonates at high surgical risk and distinguish them from those with self-limiting disease.

## Methods

### Study design and setting

We conducted a prospective observational cohort study of 67 neonates admitted between January 2021 and January 2025 to either a tertiary perinatal or surgical neonatal intensive care unit with a clinical diagnosis of suspected NEC. Following informed parental consent, all neonates underwent abdominal radiography (plain abdominal X-ray) as standard care, followed by point-of-care abdominal US within four hours of their initial X-ray. Clinical data collected included gestational age, birth weight, type of treatment (medical or surgical) and outcomes at discharge.

### Case classification

Case definitions were determined prior to study commencement. At study completion, each case was categorised as definite NEC, suspected NEC, or non-NEC. Classification was based on final clinical diagnosis, operative findings, radiographic evidence of pneumatosis intestinalis or portal venous gas, and overall clinical course. Neonates treated medically with non-specific radiographic findings were designated as having suspected NEC.

### Image acquisition and review

Radiographs and ultrasound images were de-identified, separated from clinical data, and stored for blinded analysis. Three investigators independently reviewed all X-rays classifying them as reassuring and non-reassuring based on the pre-defined criteria. **Supplementary table 1.** Disagreement in four cases was resolved by consensus. US scans were classified separately by an investigator with eight years of point-of-care abdominal US experience, also using pre-defined criteria and blinded to clinical outcomes. **Supplementary table 2.**

### Statistical analysis

Continuous variables were summarised as mean (SD) and categorical variables as frequency (percentage). Associations between imaging category (reassuring or non-reassuring) and diagnostic group (definite, suspected, or non-NEC) were examined using the χ² test. **Supplementary table 3.**

To explore whether imaging patterns could distinguish surgical from non-surgical cases, we performed two unsupervised clustering analyses: one using X-ray findings alone and another combining X-ray and US findings. Predictor matrices were constructed from complete cases and standardised as z-scores. K-means clustering (k = 2) with Euclidean distance was applied to partition neonates into two data-driven groups corresponding to lower and higher surgical risk. The choice of k = 2 was supported by inspection of the elbow plot (within-cluster sum of squares), silhouette, and gap statistics. Cluster quality was summarised by average silhouette width.^17^

The clinical concordance with the reference outcome - neonate undergone surgery (surgery: Yes/No), was evaluated using a logistic regression model with surgery as the dependent variable and cluster membership as the predictor (reporting odds ratios with 95% confidence intervals) and by comparing observed proportions of surgery between the clusters. **Supplementary table 4.**

To facilitate interpretation of the multimodal imaging model, we performed Principal component analysis (PCA) on the same scaled matrix used for clustering. We assessed the variance explained by Principal components 1 and 2 (PC1 and PC2) and calculated variable loadings to identify imaging features contributing most strongly to separation. Biplots of participant scores and variable vectors were generated to visualise feature-alignment with the cluster structure.^18^

All analyses were performed in R (version 4.3.2; R Foundation for Statistical Computing, Vienna, Austria). This study was approved by the Sydney Children’s Hospitals Network Human Research Ethics Committee (Approval ID: 2020/ETH01809).

## Results

### Cohort characteristics

Sixty-seven neonates with clinically suspected NEC were enrolled. The mean (SD) gestational age was 29.5 (4.4) weeks, and the mean birth weight was 1411 (909) grams. Fifty-nine neonates (88%) were born preterm, and 12 (18%) were growth-restricted. Thirty-three neonates (49%) underwent surgery, and seven (10%) died before discharge. **Table 1**.

**Table 1:** Baseline characteristics of infants with suspected necrotising enterocolitis (n = 67).

| Characteristic | Value |
| --- | --- |
| Gestational age (weeks), mean (SD) | 29.5 (4.4) |
| Gestational age, n (%) |  |
| Preterm ($\leq 37$ weeks) | 59 (88%) |
| Term ($>37$ weeks) | 8 (12%) |
| Birth weight (g), mean (SD) | 1411 (909) |
| IUGR, n (%) | 12 (18%) |
| Gender |  |
| Male, n (%) | 41 (61%) |
| Female, n (%) | 26 (39%) |
| Surgery, n (%) | 33 (49%) |
| Upper GI contrast study, n (%) | 10 (15%) |
| Death, n (%) | 7 (10%) |
| **Final diagnosis classification , n (%)** |  |
| **NEC** | 32 (48%) |
| **Non-NEC** | 35 (52%) |
| Total NEC cases (definite + suspected) | 32 (48%) |
| Definite NEC (Laparotomy + clinical course) | 23 (34%) |
| Suspected NEC (medical treatment only) | 9 (13%) |
| Non-NEC (other surgical diagnosis / feeding intolerance) | 35 (52%) |
| **US finding categories, n (%)** |  |
| Reassuring | 27 (40%) |
| Non-reassuring | 40 (60%) |
| **X-ray finding categories, n (%)** |  |
| Reassuring | 4 (6%) |
| Non-reassuring | 63 (94%) |

*SD=standard deviation, n=number, IUGR =intrauterine growth restriction, NEC=necrotising enterocolitis, GI=gastrointestinal*

### Final diagnostic classification

At discharge, 23 neonates (34%) were diagnosed with *definite NEC*, of whom 16 had operative confirmation. Nine (13%) were classified as *suspected NEC*, and 35 (52%) were categorised as *non-NEC*, including 18 neonates with feeding intolerance. **(Table 1**, **Figure 1)**

**Figure 1:**
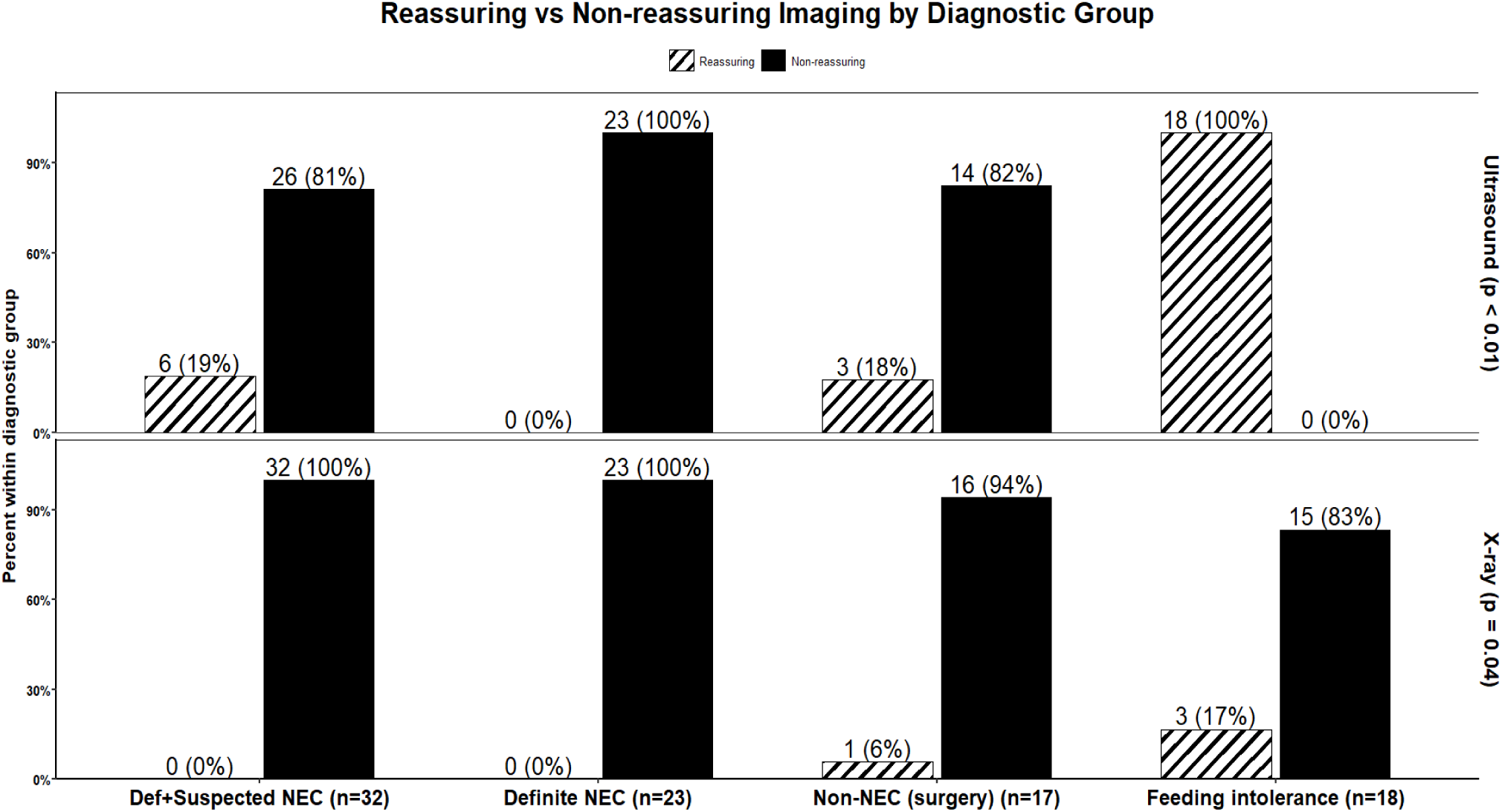
Comparison of radiograph and ultrasound classifications (reassuring vs non-reassuring) across diagnostic categories. Ultrasound was reassuring in all non-necrotising feeding intolerance cases, whereas most X-rays were non-reassuring.

### Radiographic and ultrasound finding categories

US scans were categorised as reassuring in 27 neonates (40%), non-reassuring in 40 (60%). X-rays were reassuring in only 4 infants ( 6%) and non-reassuring in 63 infants (94%). **Table 1**

Among infants with non-NEC feeding intolerance, US findings were reassuring in all cases, whereas 83% of X-rays were non-reassuring. In contrast, both imaging modalities were non-reassuring in all infants with definite NEC. **Figure 1**

### Cluster analysis

Two unsupervised k-means cluster models were derived. In the X-ray–only model, the proportion requiring surgery did not differ significantly between clusters (58.8% vs 39.4%; p = 0.11) **Figure 2**.

**Figure 2:**
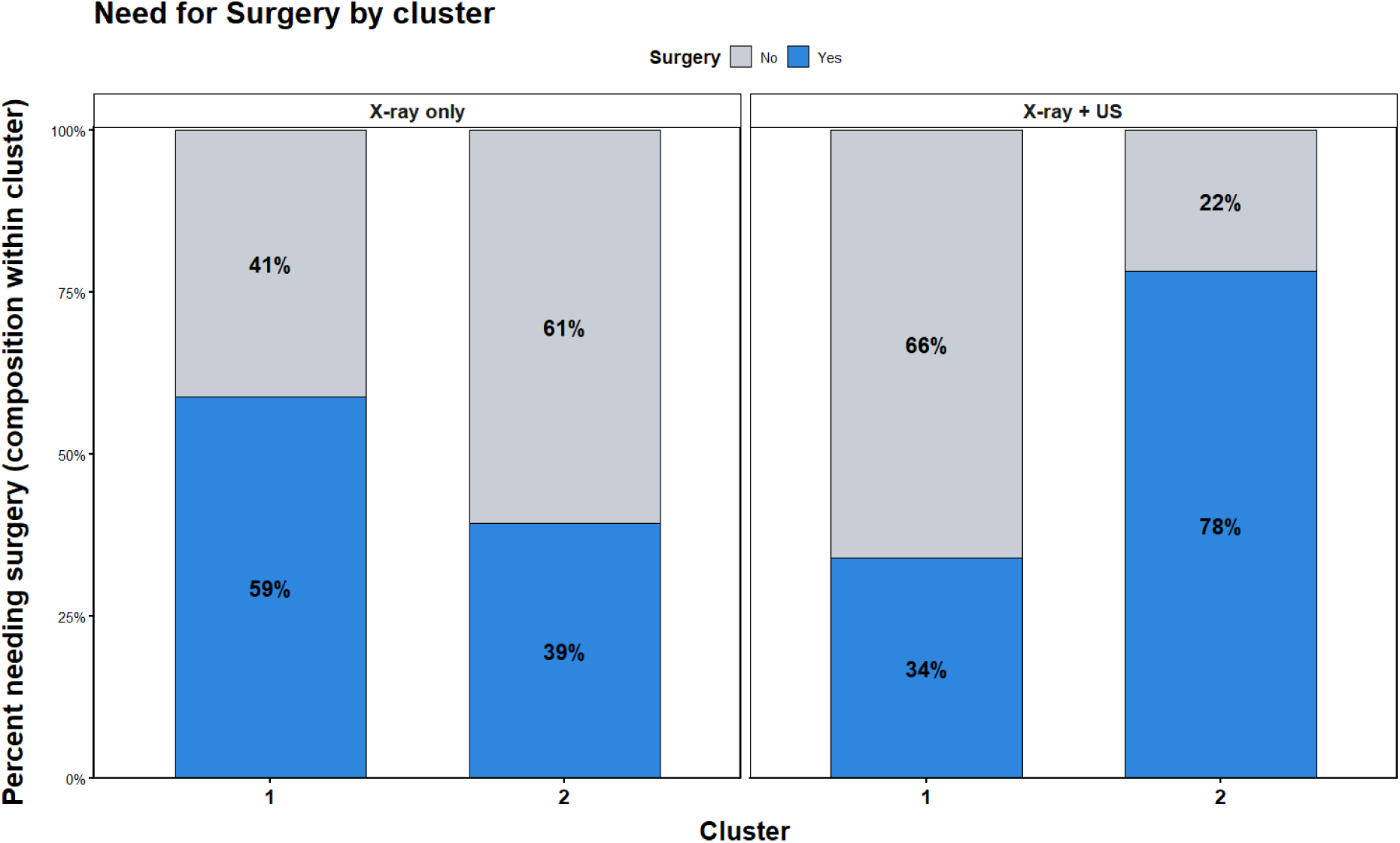
Surgical outcomes by imaging model. Each pair of bars shows the proportion of infants requiring surgery within two clusters derived from k-means analysis. X-ray–only model: surgical proportions did not differ significantly between clusters (58.8% vs 39.4%; p = 0.11) Combined X-ray + ultrasound model: clusters showed clear separation (78.3% vs 34.1%; OR 6.96, 95% CI 2.29 – 24.58), indicating improved surgical risk stratification with multimodal imaging.

In the combined X-ray and US model, clustering produced two distinct groups with markedly different surgical rates (78.3% vs 34.1%), corresponding to an odds ratio of 6.96 (95% CI 2.29–24.58).

### Principal component analysis (PCA)

PCA of the combined imaging dataset showed that the first two components explained 70.1% of the total variance. The biplot revealed clear separation between neonates who underwent surgery and those managed medically. Complex ascites, absent peristalsis, abnormal bowel perfusion, and abnormal bowel gas pattern were the features, most strongly associated with the surgical cluster. The resulting biplot and loading plot are shown in **Figure 3**. The loading plot demonstrates the contribution of each radiological finding (variable) to these components **Table 2**.

**Figure 3:**
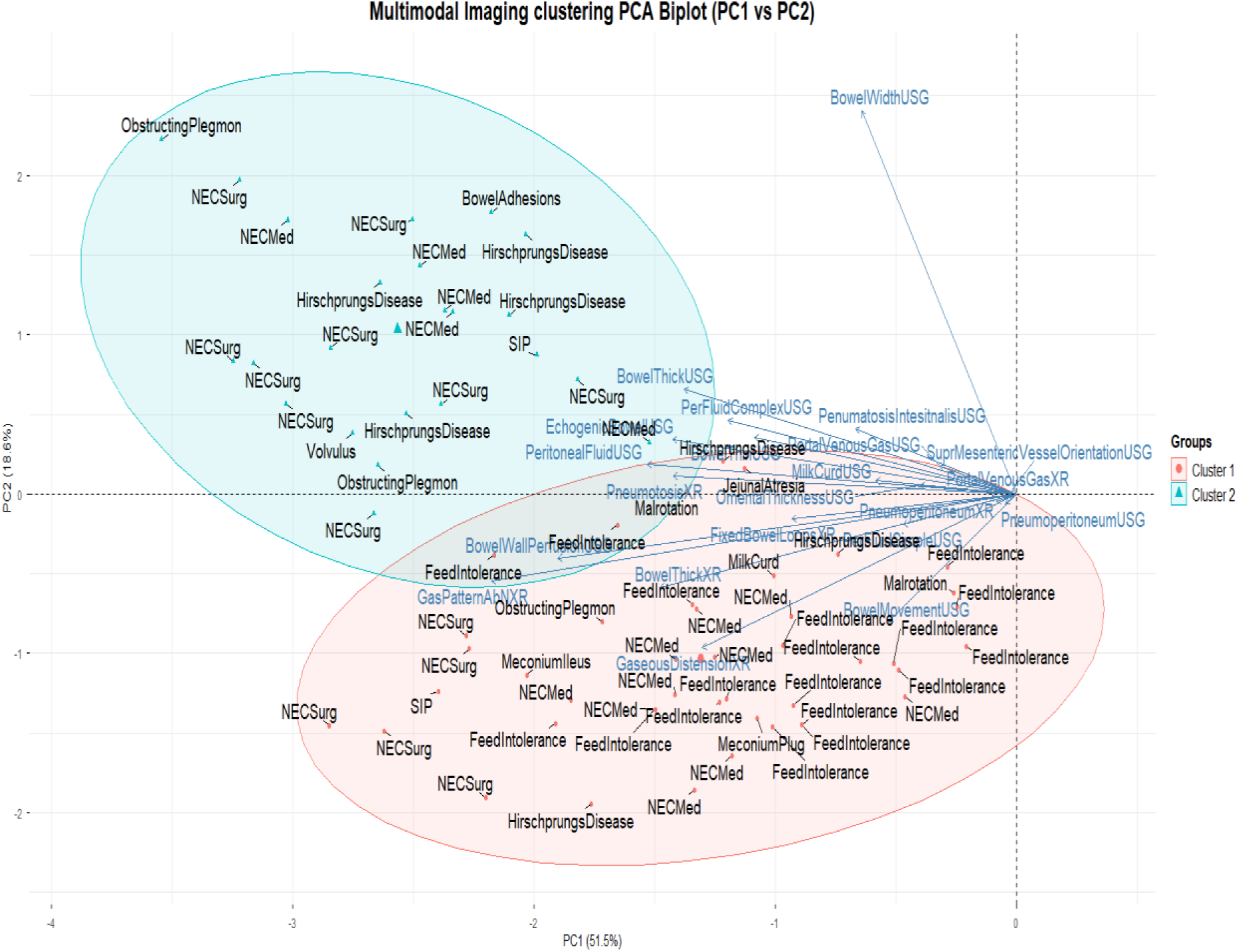
PCA biplot segregation of cases based on both imaging findings into two-clusters (cluster 1 circle, cluster 2 triangle).

**Table 2:** Principal component analysis loadings for combined X-ray and ultrasound features (n = 67).

| PC1 |  |  | PC2 |  |  |
| --- | --- | --- | --- | --- | --- |
| PC1 Rank | PC1<br>Abnormal findings on X-Ray and US to differentiate between need for surgery | PC1 Loading | PC2 Rank | PC2<br>Abnormal findings on X-Ray and US to differentiate between need for surgery | PC2 Loading |
| 1 | Abnormal bowel gas pattern on X-ray | -0.427 | 1 | Bowel wall width on US | 0.785 |
| 2 | Bowel wall perfusion abnormalities on US | -0.373 | 2 | Gaseous distention on X-Ray | -0.315 |
| 3 | Peritoneal fluid present on US | -0.300 | 3 | Loss of Bowel movement on US | -0.263 |
| 4 | Bowel wall thickening on X-Ray | -0.290 | 4 | Bowel wall thickening on US | 0.215 |
| 5 | Echogenic bowel on US | -0.280 | 5 | Bowel thickening on X-Ray | -0.191 |
| 6 | Pneumatosis on X-ray | -0.279 | 6 | Abnormal gas pattern on X-Ray | -0.178 |
| 7 | Bowel thickening on US | -0.271 | 7 | Complex peritoneal fluid collection on US | 0.150 |
| 8 | Gaseous distension on X-Ray | -0.255 | 8 | Pneumatosis Intestinalis on US | 0.134 |
| 9 | Complex peritoneal fluid collection on US | -0.234 | 9 | Bowel wall perfusion Abnormalities on US | -0.131 |
| 10 | Bowel wall thinning on US | -0.213 | 10 | Bowel wall thinning US | 0.117 |

*US=ultrasound*
*Positive loadings indicate association with the high-risk (surgical) cluster.*
*Features with higher absolute loadings contribute most to component separation.*

## Discussion

This prospective cohort study demonstrates that combining abdominal US with radiography improves surgical risk stratification in infants with suspected NEC compared with radiography alone. Multimodal imaging produced distinct, data-driven clusters that corresponded closely to surgical outcomes, whereas X-ray alone failed to achieve a clear separation. A reassuring US in clinically suspected NEC cases, particularly those later diagnosed with non-necrotising conditions such as feeding intolerance, consistently aligned with a low likelihood of surgery. These findings suggest that incorporating US into the early clinical evaluation of neonates with suspected NEC may enhance diagnostic confidence and inform safer clinical decision-making.

For neonates with suspected NEC, radiography remains an essential first-line investigation, particularly for detecting pneumoperitoneum and bowel gas distribution. However, plain abdominal X-rays offer limited insight into bowel perfusion or viability.^4^ Its low specificity was evident in this cohort, where most X-rays were categorised as non-reassuring despite substantial heterogeneity in disease severity. US provided synergistic diagnostic value by providing real-time assessment of bowel wall perfusion, thickness, and peristalsis, and the presence of complex ascites, a feature that correlates with surgical intervention.^19–22^ The ability of US to identify these early pathophysiological changes in NEC, may explain its superior discriminative diagnostic performance.

Our PCA report supports previous work identifying complex fluid, absent peristalsis, and reduced perfusion in NEC, as markers of severe disease.^23^ These findings align with those reported by Cuna et al. ^23^ and Lazow et al. ^24^, as US features predictive of surgery and mortality in NEC. Interestingly, the association between portal venous gas and NEC disease severity, remains debated. While Chen and colleagues ^22^ reported a strong link of the presence of portal venous gas to the need for surgery in NEC, other series have shown less consistent results, suggesting that the timing of performing the US scan and the operator experience, may influence its interpretation.

Beyond diagnostic accuracy, the integration of US into routine assessment has practical implications.^15^ A reassuring US may help avoid unnecessary transfers, reduce exposure to broad-spectrum antibiotics, and support earlier re-feeding in selected infants. Conversely, early identification of high-risk imaging features can prompt timely surgical consultation and improve preparedness for deterioration.^22^ Establishing standardised imaging criteria and scoring systems across centres could facilitate more consistent communication between neonatologists, radiologists, and surgeons.^15^

The study design incorporated several features that strengthen its validity. Imaging was performed prospectively within a defined time window, and analyses were conducted using masked data to reduce bias. Unsupervised clustering provided an objective, data-driven framework for exploring imaging patterns without imposing clinical assumptions. The addition of PCA enhanced interpretability by identifying features most responsible for surgical-risk separation.

Several limitations should be acknowledged. This was a two-centre study with a modest sample size, limiting generalisability. US interpretation was performed by a single experienced operator, and inter-rater variability was not assessed.

Although cluster membership was associated with surgical intervention, the model did not incorporate clinical or laboratory data that may further improve prediction accuracy. Future studies should validate these findings across multiple sites, assess inter-observer agreement, and explore integration of multimodal imaging with biochemical and physiological parameters.

### Conclusions

Combining abdominal ultrasound with radiography improved the identification of neonates at high surgical risk from NEC. The ultrasound provided valuable dynamic functional information, that complemented static radiographic findings and enhanced diagnostic precision. Broader adoption of multimodal imaging and standardised reporting may support a more accurate diagnosis, targeted management, and improved clinical outcomes for neonates with NEC.

## Data Availability

All data produced in the present study are available upon reasonable request to the authors

