## Supplementary Tables for "Combining Abdominal Ultrasound and Radiography for Surgical Risk Prediction in Necrotising Enterocolitis: A Prospective Cohort Study"

### SUPPLEMENTARY MATERIAL:

**Supplementary Table 1:** X-Ray findings were classified as reassuring (R) and non-Reassuring (NR) based on the following pre-defined criteria[4]

| Reassuring X-Ray | Non-reassuring X-Ray |
|---|---|
| Normal bowel gas pattern<br>No Pneumatosis intestinalis<br>No intraperitoneal gas<br>No Portal Venous Gas | Paucity of bowel gas<br>Bowel wall thickness with bowel distension<br>Gasless abdomen<br>Pneumatosis intestinalis<br>Portal venous gas<br>Intraperitoneal gas<br>Significant abnormal bowel gas pattern<br>Mild distention with bubbly gas / stool like appearance / Mild gaseous distention / absent gas in the rectum |

**Supplementary Table 2:** Ultrasound findings were classified as reassuring (R) and non-Reassuring (NR) based on the following pre-defined criteria[1,4,9,15,23]

| Reassuring US | Non-reassuring US |
|---|---|
| Normal bowel wall vascularity<br>No Pneumatosis intestinalis<br>No intraperitoneal gas<br>No Portal Venous Gas<br>A small / insignificant amount of simple fluid collection<br>Normal peristalsis defined as >10 per minute<br>Normal bowel wall thickness, defined as bowel wall measurement between 1mm-2mm | Abnormal bowel wall vascularity<br>Bowel wall thickness >2mm<br>Bowel wall thickness <1mm with loss of peristalsis<br>Pneumatosis intestinalis<br>Portal Venous Gas<br>Intraperitoneal gas<br>Loss of peristalsis<br>Complex ascites |

US = ultrasound

**Supplementary Table 3:** Ultrasound and X-ray categories v/s Final outcome (n=67)

| **Final diagnosis** | **Definite NEC + suspected NEC (n=32)** | **Definite NEC (n=23)** | **Non-NEC confirmed on surgery (n=17)** | **Non-NEC Feeding intolerance (n=18)** | **p-value** |
|---|---|---|---|---|---|
| US categories, n (%)<br>Reassuring<br>Non-reassuring | 6 (19%)<br>26 (81%) | 0 (0%)<br>23 (100%) | 3 (18%)<br>14 (82%) | 18 (100%)<br>0 (0%) | <0.01 |
| X-ray categories, n (%)<br>Reassuring<br>Non-reassuring | 0 (0%)<br>32 (100%) | 0 (0%)<br>23 (100%) | 1 (6%)<br>16 (94%) | 3 (17%)<br>15 (83%) | 0.04 |

**Supplementary Table 4:** Unsupervised clustering into two groups (C1, C2) using X-ray findings only and combined X-ray + ultrasound findings

| **Imaging categories** | **Odds Ratio (95% CI) (C2 vs C1)** | **p-value** | **Rate of Surgery C1** | **Rate of Surgery C2** |
|---|---|---|---|---|
| X-ray only | 0.45 (0.17–1.20) | 0.114 | 58.8% | 39.4% |
| Combined using X-ray + US | 6.96 (2.29 – 24.58) | 0.001 | 34.1% | 78.3% |

US = ultrasound; OR = odds ratio; CI = confidence interval
